# Prevalence, Risk Factors, Hesitancy, and Attitudes Toward Vaccination Against Reproductive Tract Infections Among Women in an Urban Setting: A Cross-Sectional Survey Study

**DOI:** 10.1101/2025.02.13.25322217

**Authors:** Sunita K. Yadav, Priya Bhardwaj, Ravi Kant, Joyeta Ghosh, Anita Garg Mangla, Sumathi Muralidhar, Daman Saluja, Rita Singh, Aleksandra E. Sikora, Jyoti Taneja

**Author notes:** **Corresponding Authors: Dr. Jyoti Taneja**, Department of Zoology, Daulat Ram College, University of Delhi, Delhi, India,;, **Professor Aleksandra E. Sikora**, Department of Pharmaceutical Sciences, College of Pharmacy, Oregon State University, Corvallis, Oregon, USA. These authors contributed equally to this work (Sunita K. Yadav, Priya Bhardwaj).

## Abstract

**Background:** Reproductive tract infections (RTIs) including sexually transmitted infections (STIs), endogenous and iatrogenic infections are a major health concern globally, particularly among young women in developing nations. If untreated, they can lead to infertility, increased HIV susceptibility, and cervical cancer. This study examines RTI symptoms prevalence, risk factors, and hesitancy toward STI vaccination.

**Methods:** A cross-sectional study was conducted among 1,920 urban women (predominantly aged 18-25) in Delhi/NCR using a structured questionnaire. Data on demographics, reproductive health, RTI symptoms, and vaccine awareness were analyzed using SPSS 20. Chi-square tests, Fisher’s exact tests, and Bonferroni corrections were applied, with multinomial logistic regression identifying RTI risk predictors.

**Results:** Among respondents, 1,240 (64.6%) reported RTI symptoms, with 1,029 (83%) experiencing vaginal discharge and 974 (78.5%) vulval itching. Significant risk factors included early menarche, irregular menstrual cycles, poor menstrual hygiene, and prior RTI medication use. Additionally, 1,033 (53.8%) were hesitant about STI vaccination, citing safety concerns (36%, 372) and cost (18.6%, 192). However, 826 (43%) reported that healthcare provider recommendations positively influenced their decision to vaccinate.

**Conclusion:** The high prevalence of RTI symptoms highlights the urgent need for improved awareness, accessible healthcare services, and stronger vaccination promotion efforts to enhance RTI prevention and management.

## 1. Introduction

Reproductive Tract Infections (RTIs) refer to infections affecting the reproductive system in both men and women. These infections are broadly categorized into three types: sexually transmitted infections (STIs), endogenous infections, and iatrogenic infections. RTI are a major public health concern, particularly for women, as they can significantly impact reproductive health, pregnancy outcomes, and overall well-being [1–3]. Often described as a “silent” pandemic, RTI have far-reaching consequences not only for women’s sexual and reproductive health but also for their families and communities worldwide. Women of reproductive age are particularly vulnerable, especially during pregnancy, menstruation, and childbirth [1–4].

Common symptoms of RTI include lower abdominal pain, backache, abnormal vaginal discharge, vulvar itching, and genital ulcers [2, 3, 5]. Timely treatment is crucial, as untreated RTI can lead to severe complications such as ectopic pregnancy, infertility, cancer, neonatal mortality, and an increased risk of HIV transmission [3, 6–9]. Despite these risks, RTI remain widely underreported in developing countries due to stigma, low awareness, and inadequate healthcare services [3, 10]. In India alone, approximately 6% of the adult population is estimated to have one or more RTI/STIs, contributing to an annual burden of 30– 35 million cases [11].

The World Health Organization (WHO) estimates that approximately 374 million new cases of curable STIs occur annually among individuals aged 15–49 years, including gonorrhea, chlamydia, syphilis, and trichomoniasis [3, 10]. However, these estimates exclude viral STIs such as human immunodeficiency virus (HIV), hepatitis B, genital warts, and genital herpes [12]. Additionally, bacterial vaginosis (BV) has emerged as a significant RTI, characterized by an imbalance in the vaginal microbiome that increases susceptibility to other infections [13]. Given the overwhelming prevalence of RTIs and STIs, strengthening epidemiological data collection is essential for effective management and control [3]. To address this, WHO recommends four key surveillance methods at the country level: case reporting, etiological analysis, monitoring of antimicrobial resistance, and prevalence assessment [3, 14]. In resource-limited settings where diagnostic facilities are inaccessible, syndromic case management-based on identifying signs and symptoms rather than laboratory diagnoses—remains the primary approach for STI control [3, 15].

Despite advancements in healthcare, the persistently high prevalence of RTI underscores the need for evidence-based interventions targeting key factors driving disease transmission. Vaccines offer a promising preventive strategy, as demonstrated by the success of human papillomavirus (HPV) and hepatitis B vaccines, which have significantly reduced genital tract infections and related complications [3, 16]. Ongoing research is exploring new STI vaccines, including a chlamydia vaccine currently in Phase I clinical trials [17]. While no licensed gonorrhea vaccine exists, *Neisseria meningitidis* serogroup B outer membrane vesicle-based vaccines, such as MenB-4C (Bexsero®), may offer cross-protection against gonorrhea, with several clinical trials assessing their efficacy in preventing *N. gonorrhoeae* infections [18]. However, despite their potential in reducing STI prevalence, public acceptance of STI vaccines remains a major challenge due to misinformation, safety concerns, and cultural resistance. Understanding public attitudes, barriers, and misconceptions regarding STI vaccines through surveys on vaccine hesitancy is essential. Accordingly, this study aims to determine the prevalence of RTI symptoms among urban women in Delhi, identify key risk factors associated with RTI, and assess vaccine hesitancy by exploring barriers and motivating factors influencing STI vaccine acceptance. Our findings provide insights that will help public health officials, policymakers, and researchers design targeted interventions to improve vaccine uptake and combat misinformation, ultimately contributing to the broader goal of reducing the burden of RTIs and STIs.

## 2. Materials and Methods

### 2.1. Data Source

A cross-sectional survey study was conducted among urban women in Delhi/NCR from August 2024 to October 2024. The survey questionnaire, designed and validated with input from experts and senior gynecologists specializing in RTI treatment, was administered via Google Forms. This pre-validated questionnaire covered various aspects, including demographics, reproductive health, menstrual hygiene, contraceptive awareness, comorbidities among women and their family members, and attitudes toward acceptance well as hesitancy regarding RTI vaccination.

The survey was conducted online using Google Forms and followed the snowball sampling method to enhance randomness. The survey link was disseminated via WhatsApp and email to urban women in Delhi/NCR. Participants were also encouraged to share the questionnaire further to maximize participation.

### 2.2. Inclusion criterion

The online survey was administered to women aged 18 and above in Delhi. Informed consent was obtained from all participants. Only those who provided complete information in the Google Form were included in the study.

### 2.3. Exclusion criterion

Participants younger than 18 years were excluded from the study. Additionally, those who did not provide complete information in the Google Form were also excluded.

### 2.4. Study Design and Methodology

This study adhered to the Strengthening the Reporting of Observational Studies in Epidemiology (STROBE) guidelines to ensure transparent and standardized reporting. A total of 1,935 responses were collected from women residing in Delhi via an online Google survey form. Of these, 15 responses were excluded due to participants being under 18 years old or providing incomplete details, leaving 1,920 responses for analysis.

### 2.5. Participant Classification

Among the 1,920 analyzed responses, 1,241 (64.6%) were categorized as having RTI symptoms, based on self-reported symptoms suggestive of reproductive tract infections (RTIs). This group was further stratified into: 1) Low-risk RTI (*N* = 446, 23.2%) – Participants reporting two or fewer symptoms; 2) High-risk RTI (*N* = 795, 41.4%) – Participants reporting more than two symptoms.

### 2.6. Data Analysis

All responses were compiled in Microsoft Excel and analyzed using SPSS 20. The statistical methods applied included descriptive analyses for sociodemographic and categorical variables, as well as chi-square tests, Fisher’s exact tests, and Bonferroni corrections for pairwise comparisons among low-risk RTI, high-risk RTI, and RTI-absent groups, with a significance threshold of 0.0167. Additionally, multinomial logistic regression analysis was conducted to identify risk factors for RTI among urban women, using a 5% level of significance for all statistical comparisons.

### 2.7. Visualization and Illustrations

Graphs were generated using GraphPadPrism10, and illustrations were created using BioRender.com to visually represent the study findings.

## 3. Results

### 3.1. Demographic Profile of Participants

A flow diagram detailing participant inclusion, exclusion, and classification is presented in **Figure 1**, following STROBE guidelines. The demographic profile of respondents is presented in **Table 1**. The majority of participants (94.6%) were between 18 and 25 years old, while the remaining 5.4% were aged 26 to 50.

**Figure 1.**
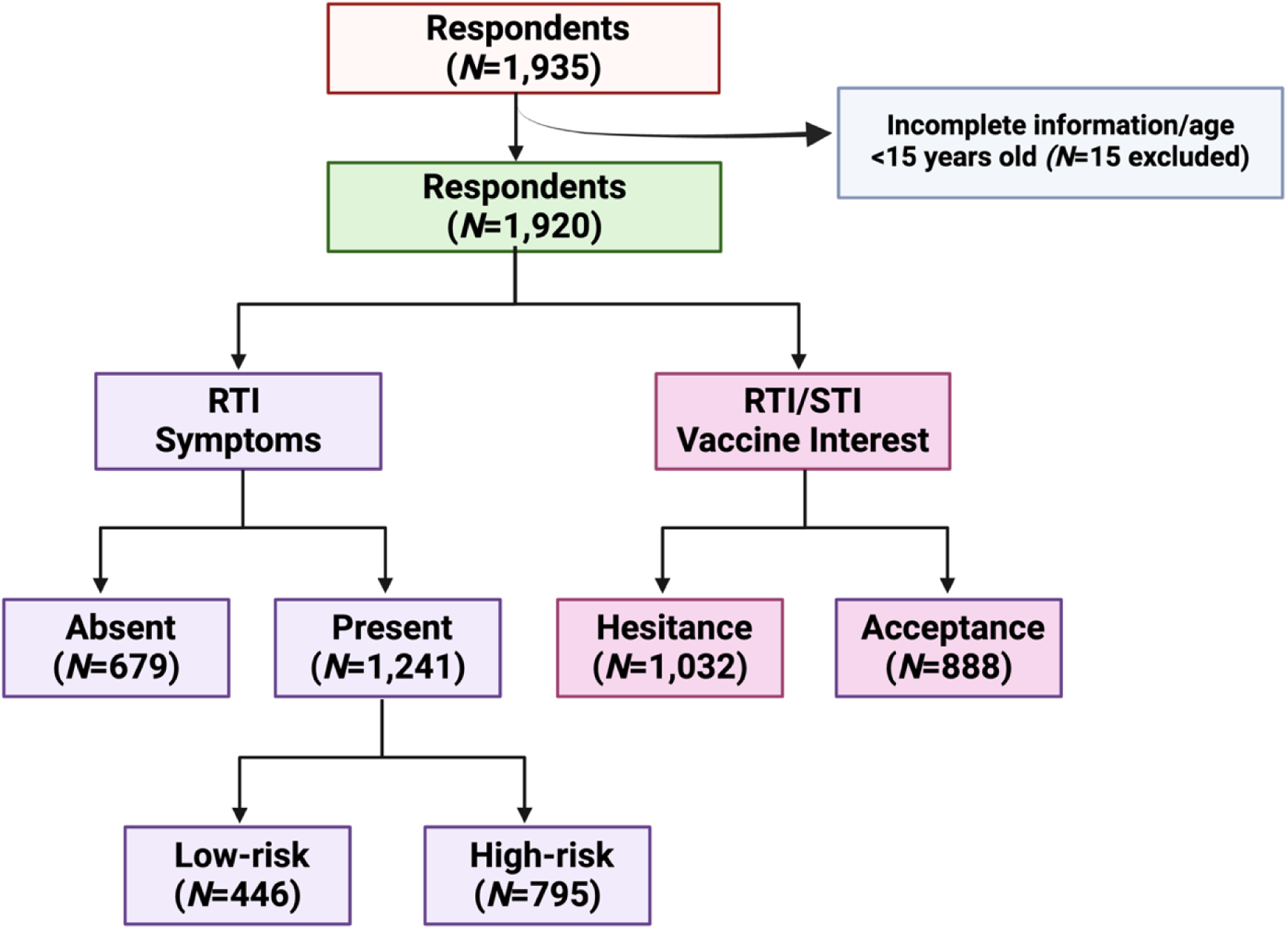
STROBE Flow Diagram of Participant Selection and Classification. This flow diagram illustrates the selection process and classification of study participants. A total of 1,935 respondents were initially screened. After excluding 15 individuals due to incomplete information or being under 18 years old, 1,920 respondents were included in the final analysis. Participants were categorized based on the presence of reproductive tract infections (RTIs) symptoms and their interest in RTIs/sexually transmitted infections (STIs) vaccination.

**Table 1.**
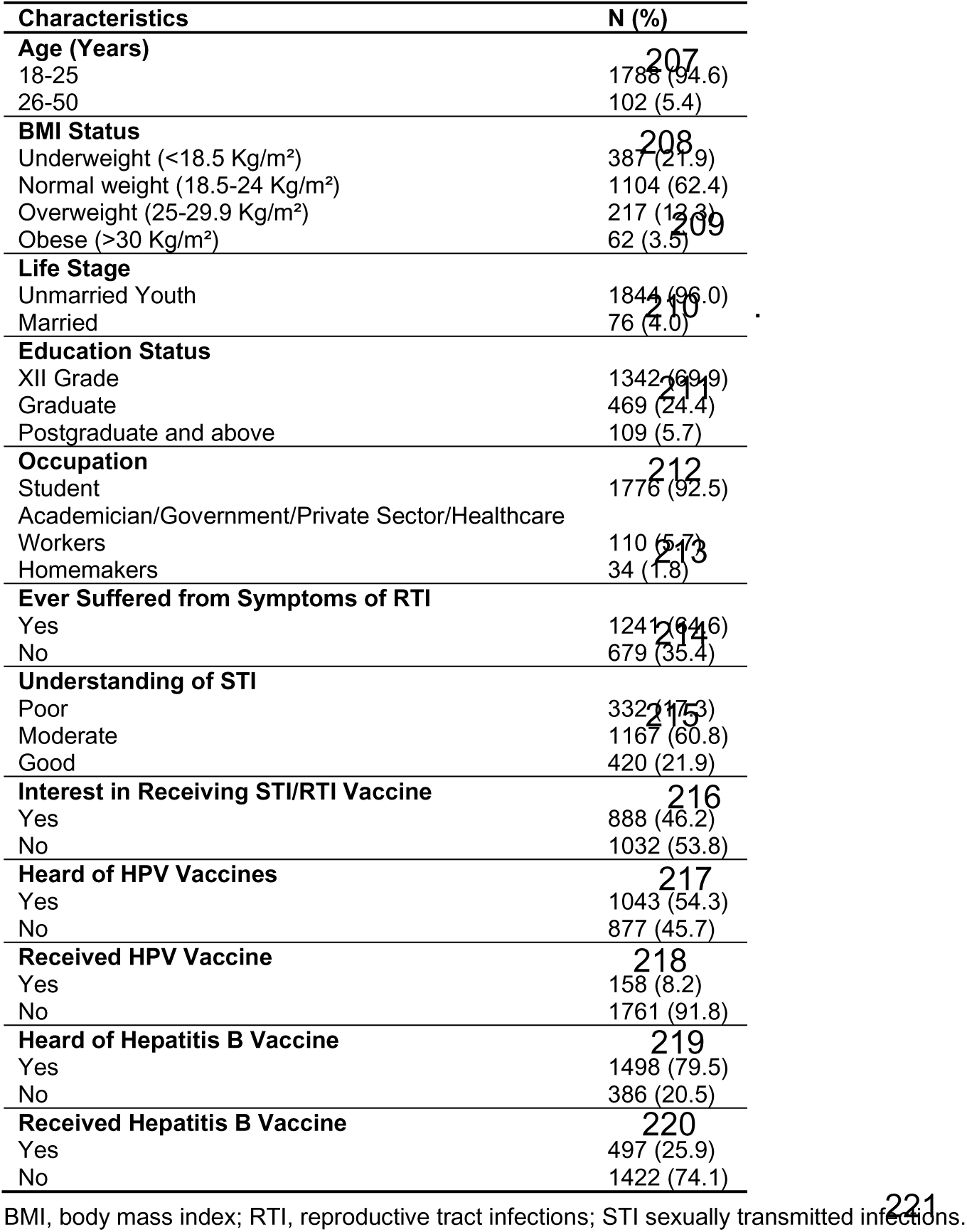
Sociodemographic and Health Awareness Characteristics of Study Participants (N=1920)

Among all respondents, 62.4% had a normal BMI (body mass index), while 21.9% were underweight, 12.3% were overweight, and 3.2% were obese.

Regarding marital status, 96% of respondents were unmarried, while the remaining 4% were married. In terms of education, 69.9% had completed up to the twelfth grade, followed by graduates (24.4%) and postgraduates or higher (5.7%). Most of participants (92.5%) were students, while 5.7% were academicians and 1.8% were homemakers.

Among the total respondents, 64.6% reported experiencing symptoms suggestive of RTI, while 35.4% did not. Therefore, the prevalence of RTI among women in Delhi was found to be 64.6%. When asked about their understanding of STIs, 60.8% had a “moderate understanding,” 21.9% had a “good understanding,” and 17.3% had a “poor understanding.“

Regarding their willingness to receive vaccines against STIs, 53.8% of participants were hesitant, while 46.2% were willing to get vaccinated. Additionally, 45.7% of participants were unaware of the HPV (human papillomavirus) vaccine, and 20.9% were unaware of the hepatitis B vaccine. However, 8.2% had already received the HPV vaccine, and 25.9% had been vaccinated against hepatitis B. Participants were also asked about the age at which they received these vaccines. Among those who had received the HPV vaccine (*N*=158, 8.2%), only 89 (56.3%) reported their age at the time of vaccination. Most of them (*N*=70) had received the vaccine between the ages of 11 and 18. Similarly, among the 497 participants who had received the hepatitis B vaccine, 276 (55.5%) reported their age at vaccination. The minimum reported age for receiving the hepatitis B vaccine was below 1 year (*N*=276), while the maximum was 30 years (*N*=76). The majority (*N*=200) had received the vaccine before turning 1 year old

### 3.2. Prevalence of Reproductive Tract Infections (RTIs)

The prevalence of RTI was assessed using the WHO syndromic method guidelines. During the study, participants were asked whether they had experienced any symptoms listed in the questionnaire. Among all participants, 64.6% (*N*=1241) reported at least one symptom suggestive of RTI. Of these, 23.2% (*N*=446) reported two or fewer symptoms and were categorized as the low-risk RTI group. The remaining 41.4% (*N*=795) reported more than two symptoms and were classified as the high-risk RTI group (**Figure 1**).

#### 3.2.1. Frequency of Symptoms Suggestive of RTI

The symptoms reported by participants in both low- and high-risk RTI groups are shown in **Figure 2**. Among high-risk RTI participants, the most commonly experienced symptoms were vaginal discharge (83%), followed by vulvar itching (78.5%), lower abdominal pain (75.8%), backache (65.7%), urinary tract infections (24.4%), perianal pain (20.6%), dysuria (17.9%), polyuria (14.6%), and abnormal vaginal growth (4.8%). In contrast, the most prevalent symptoms among low-risk RTI participants were lower abdominal pain (58.5%) and backache (58.1%).

**Figure 2.**
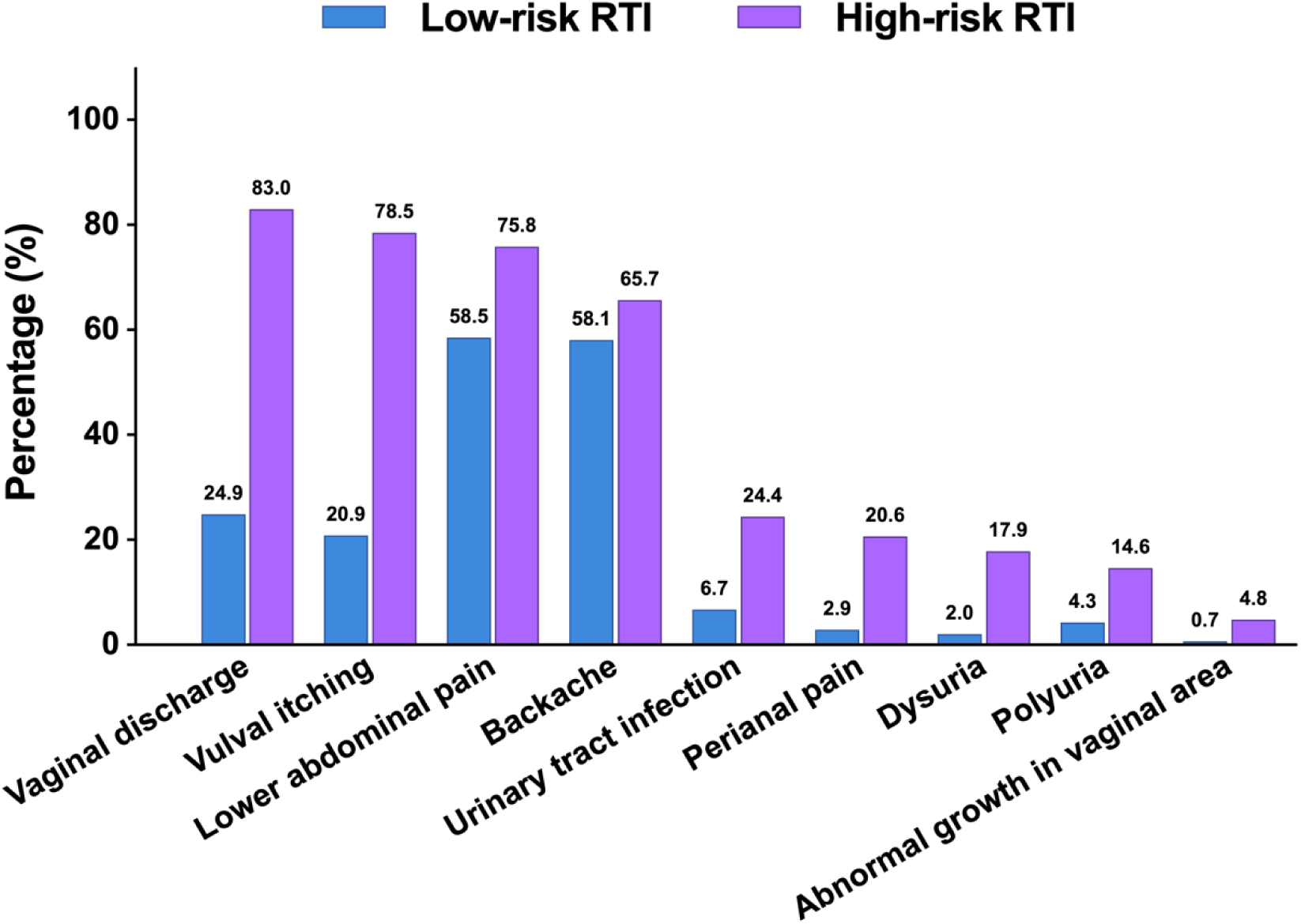
Distribution of Symptoms Among Low- and High-Risk RTI Groups. This bar chart presents the prevalence of symptoms associated with reproductive tract infections (RTI) among participants classified as low-risk and high-risk. The x-axis represents different RTI symptoms, while the y-axis shows the percentage of participants experiencing each symptom. The percentage for each symptom is displayed above its corresponding bar in the group.

#### 3.2.2. Gynecological Diagnostic History of Participants Belonging to Low- and High-Risk RTI

The gynecological diagnostic history of participants from both low- and high-risk RTI groups was examined. Participants were asked whether they had ever been diagnosed with an RTI. Among the total participants (*N*=1,241), approximately 5% (*N*=57) had a documented history of gynecological diagnoses.

The most commonly diagnosed gynecological condition was pelvic inflammatory disease (affecting the endometrium, fallopian tubes, and ovaries), accounting for 39.5% of cases across both groups (**Supplementary Figure S1**). However, vaginitis was notably more prevalent in the high-risk RTI group (37.2%) compared to the low-risk group (28.6%).

Other reproductive infections, such as vulval and Bartholin gland infections, were slightly more frequent in the low-risk RTI group (21.4%) than in the high-risk RTI group (16.8%). Cervicitis was the least reported RTI, with similar rates in both groups (6.9% and 7.1%, respectively; **Supplementary Figure S1**).

### 3.3. Association of Sociodemographic, Reproductive, Hygiene, Contraceptive, and Vaccination Factors with RTI Risk Levels

#### 3.3.1. Sociodemographic Factors

In this study, sociodemographic variables were not found to be associated with low- or high-risk RTI (**Table 2**). The majority of participants (over 93%) across all three groups were between 18 and 25 years old. Most participants had a normal weight, were unmarried, and had at least a twelfth-grade education.

**Table 2.** Association of Demographic Factors with Symptoms of Reproductive Tract Infections in Urban Women.

| Variables | RTI absent<br>N=679 (35.4%) | Low-risk RTI<br>N=446 (23.2%) | High-risk RTI<br>N=795<br>(41.4%) | Low-risk RTI vs<br>RTI absent<br>$\chi^2$<br>P-value | High-risk RTI<br>vs RTI absent<br>$\chi^2$<br>P-value | Low-risk RTI<br>vs high-risk RTI<br>$\chi^2$<br>P-value |
| --- | --- | --- | --- | --- | --- | --- |
| **Age (years)** |  |  |  |  |  |  |
| 18-25 | 628 (93.7%) | 418 (94.8%) | 742 (95.2%) | 4.8399 | 4.4130 | 8.9883 |
| 26-50 | 42 (6.3%) | 23 (5.2%) | 37 (4.8%) | 0.1839 | 0.2202 | 0.0294* |
| **BMI Status** |  |  |  |  |  |  |
| Underweight (<18.5 Kg/m<sup>2</sup>) | 143 (23.0%) | 90 (22.3%) | 154 (20.7%) | 0.2097 | 2.1187 | 2.1187 |
| Normal weight (18.5-24 Kg/m<sup>2</sup>) | 386 (62.2%) | 253 (62.6%) | 465 (62.4%) | 0.9760 | 0.5481 | 0.5481 |
| Overweight (25-29.9 Kg/m<sup>2</sup>) | 67 (10.8%) | 46 (11.4%) | 104 (14.0%) |  |  |  |
| Obese (>30 Kg/m<sup>2</sup>) | 25 (4.0%) | 15 (3.7%) | 22 (2.9%) |  |  |  |
| **Marital Status** |  |  |  |  |  |  |
| Unmarried youth | 651 (95.9%) | 426 (95.5%) | 767 (96.5%) | 0.9315 | 1.5702 | 3.1439 |
| Married | 28 (4.1%) | 20 (4.5%) | 28 (3.5%) | 0.8178 | 0.6662 | 0.3700 |
| **Education Status** |  |  |  |  |  |  |
| XII grade | 461 (67.9%) | 307 (68.8%) | 574 (72.2%) | 2.3482 | 3.4486 | 5.3175 |
| Graduate | 179 (26.4%) | 105 (23.5%) | 185 (23.3%) | 0.3091 | 0.1783 | 0.0700* |
| Postgraduate and above | 39 (5.7%) | 34 (7.6%) | 36 (4.5%) |  |  |  |
| **Occupation** |  |  |  |  |  |  |
| Student | 626 (92.2%) | 408 (91.5%) | 742 (93.3%) | 1.8578 | 3.8842 | 2.8944 |
| Academician/Government/ Private<br>company/Healthcare workers | 38 (5.6%) | 29 (6.5%) | 43 (5.4%) | 0.7619 | 0.4219 | 0.5757 |
| Home Makers | 15 (2.2%) | 9 (2.0%) | 10 (1.3%) |  |  |  |

\*98.33% confidence intervals were calculated after correction to significance level using Bonferroni method. BMI, body mass index; RTI, reproductive tract infections.

#### 3.3.2. Reproductive, Menstrual Hygiene, and Contraceptive Awareness Factors

The age of menarche onset was significantly different between women in the high-risk RTI and RTI-absent groups. A majority (72.7%) of women in the RTI-absent group reported menarche onset between the ages of 12 and 16, compared to those in the high-risk RTI group. Notably, menstrual status and cycle length varied significantly (*P*<0.001) between women in the low- and high-risk RTI groups. Women in the high-risk RTI group exhibited a significantly higher prevalence of irregular menstrual cycles (19.9%) and prolonged menstrual cycles (>45 days) compared to those in the low-risk RTI group.

Additionally, women in the high-risk RTI group differed significantly from those in the RTI-absent group regarding diagnoses of reproductive tract diseases and RTI/STI (**Table 3**). Women in both the low- and high-risk RTI groups also showed significant differences from the RTI-absent group regarding prior use of medication for RTI symptoms within the last three months.

**Table 3.** Association Between Menstrual Health, Contraceptive Awareness, and Symptoms Suggestive of Reproductive Tract Infection in Urban Women.

| Variables | RTI Absent<br>(N=679,<br>35.4%) | Low-Risk RTI<br>(N=446,<br>23.2%) | High-Risk RTI<br>(N=795,<br>41.4%) | Low-Risk RTI vs RTI<br>Absent<br>$\chi^2$, *P*- value | High-Risk RTI vs RTI<br>Absent<br>$\chi^2$, *P* - value | Low-Risk RTI vs High-Risk<br>RTI<br>$\chi^2$, *P* - value |
| --- | --- | --- | --- | --- | --- | --- |
| **Menarche Onset (Years)** |  |  |  |  |  |  |
| <10 | 5 (0.8%) | 6 (1.4%) | 8 (1.0%) | 11.2300, 0.0105 | 16.0804, 0.0011* | 0.3508, 0.9502 |
| 10-12 | 144 (22.8%) | 135 (30.5%) | 244 (30.9%) |  |  |  |
| 12-16 | 459 (72.7%) | 294 (66.4%) | 525 (66.5%) |  |  |  |
| >16 | 23 (3.7%) | 8 (1.8%) | 13 (1.6%) |  |  |  |
| **Menstrual Cycle Status** |  |  |  |  |  |  |
| Irregular | 94 (15.2%) | 50 (11.4%) | 157 (19.9%) | 2.7060, 0.1000 | 5.0525, 0.0246 | 13.8050, 0.0002** |
| Regular | 526 (84.8%) | 387 (88.6%) | 631 (80.1%) |  |  |  |
| **Length (days)** |  |  |  |  |  |  |
| >21 to <35 | 481 (79.5%) | 346 (84.0%) | 580 (75.6%) | 3.3738, 0.1851 | 3.6822, 0.1586 | 11.2012, 0.0037** |
| >35 to <45 | 97 (16.0%) | 50 (12.1%) | 138 (18.0%) |  |  |  |
| >45 | 27 (4.5%) | 16 (3.9%) | 49 (6.4%) |  |  |  |
| **Diagnosed with RTI** |  |  |  |  |  |  |
| Yes | 11 (1.6%) | 14 (3.1%) | 43 (5.4%) | 2.2021, 0.1378 | 11.7585, 0.0006** | 2.0248, 0.1547 |
| No | 668 (98.4%) | 432 (96.9%) | 752 (94.6%) |  |  |  |
| **Diagnosed with RTI/STI** |  |  |  |  |  |  |
| Yes | 1 (0.1%) | 3 (0.7%) | 19 (2.4%) | 0.8734, 0.3500 | 12.1155, 0.0005** | 3.9028, 0.0482 |
| No | 677 (99.9%) | 443 (99.3%) | 776 (97.6%) |  |  |  |
| **Use of Medication for RTI/UTI in the Past Three Months** |  |  |  |  |  |  |
| Yes | 33 (4.9%) | 156 (35.0%) | 241 (30.3%) | 172.2306, 0.0000** | 154.8223, 0.0000** | 2.6453, 0.1039 |
| No | 645 (95.1%) | 290 (65.0%) | 554 (69.7%) |  |  |  |
| **Menstrual Hygiene** |  |  |  |  |  |  |
| Yes | 640 (94.4%) | 439 (98.4%) | 789 (99.2%) | 10.3723, 0.0013** | 28.0528, 0.0000** | 1.1283, 0.2881 |
| No | 38 (5.6%) | 7 (1.6%) | 6 (0.8%) |  |  |  |
| **Awareness of Contraceptive Methods** |  |  |  |  |  |  |
| Yes | 404 (70.9%) | 297 (78.8%) | 574 (84.7%) | 6.9646, 0.0083** | 33.8926, 0.0000** | 5.4181, 0.0199 |
| No | 166 (29.1%) | 80 (21.2%) | 104 (15.3%) |  |  |  |

\*98.33% confidence intervals were calculated after correction to significance level using Bonferroni method. BMI, body mass index; RTI, reproductive tract infection; STI, sexually transmitted infection; UTI urinary tract infection.

Regarding menstrual hygiene, the majority of women (>94%) reported maintaining proper hygiene. However, adherence was significantly higher among women in the low- and high-risk RTI groups (**Table 3**). Similarly, awareness of RTI was significantly greater (>78%) among women in these groups compared to the RTI- absent group.

#### 3.3.3. History of Co-Morbidities

When comparing comorbidities, a significantly higher percentage of participants in the high-risk RTI group (68.1%) reported having family members who had suffered or were suffering from a comorbidity, compared to those in the low-risk (58.5%) and RTI-absent (51.8%) groups (**Table 4**). However, the percentage of participants with medical conditions was similar across all three groups, ranging from 22.4% to 25.7%.

**Table 4:** History of Comorbidities Associated with Symptoms Suggestive of Reproductive Tract Infections Among Women.

| Variables | RTI absent<br>N=679<br>(35.4%) | Low risk RTI<br>N=446<br>(23.2%) | High risk<br>RTI<br>N=795<br>(41.4%) | Low risk<br>RTI<br>vs<br>RTI absent<br>$\chi^2$<br>P-value | High risk<br>RTI<br>vs<br>RTI absent<br>$\chi^2$<br>P-value | Low risk RTI<br>vs<br>High risk<br>RTI<br>$\chi^2$<br>P-value |
| --- | --- | --- | --- | --- | --- | --- |
| **Family history of comorbidities** |  |  |  |  |  |  |
| Yes | 352 (51.8%) | 261 (58.5%) | 541 (68.1%) | 4.5770 | 39.6180 | 10.9379 |
| No | 327 (48.2) | 185 (41.5%) | 254 (31.9%) | 0.0324 | 0.0000** | 0.0009** |
| **Existing medical conditions** |  |  |  |  |  |  |
| Yes | 164 (24.8%) | 96 (22.4%) | 197 (25.7%) | 0.7194 | 0.1009 | 1.4527 |
| No | 497 (75.2%) | 333 (77.6%) | 570 (74.3%) | 0.3964 | 0.7507 | 0.2281 |

\*98.33% confidence intervals were calculated after correction to significance level using Bonferroni method. RTI, reproductive tract infection.

Among the comorbidities reported in family members or relatives, diabetes was the most common, followed by hypertension, cancer, cardiovascular diseases,

PCOS, endometriosis, and thyroid disorders (**Figure 3**). Asthma was the least reported comorbidity and was observed only in the low- and high-risk RTI groups. Similarly, when comparing medical conditions among participants, hormonal imbalances and PCOS were the most prevalent across all groups (**Figure 4**). However, the prevalence of these conditions was significantly higher in the high-risk RTI group (70.6% and 45.7%, respectively) than in the low-risk RTI (44.8% and 32.3%) and RTI-absent (25% and 19.5%) groups. Other reported medical conditions included COVID-19, thyroid disorders, and tuberculosis across all three groups. Additionally, 5.7% of participants reported other conditions such as skin problems, hypertension, and asthma. Infertility was the least reported medical condition and was found only among participants in the low- and high-risk RTI groups.

**Figure 3.**
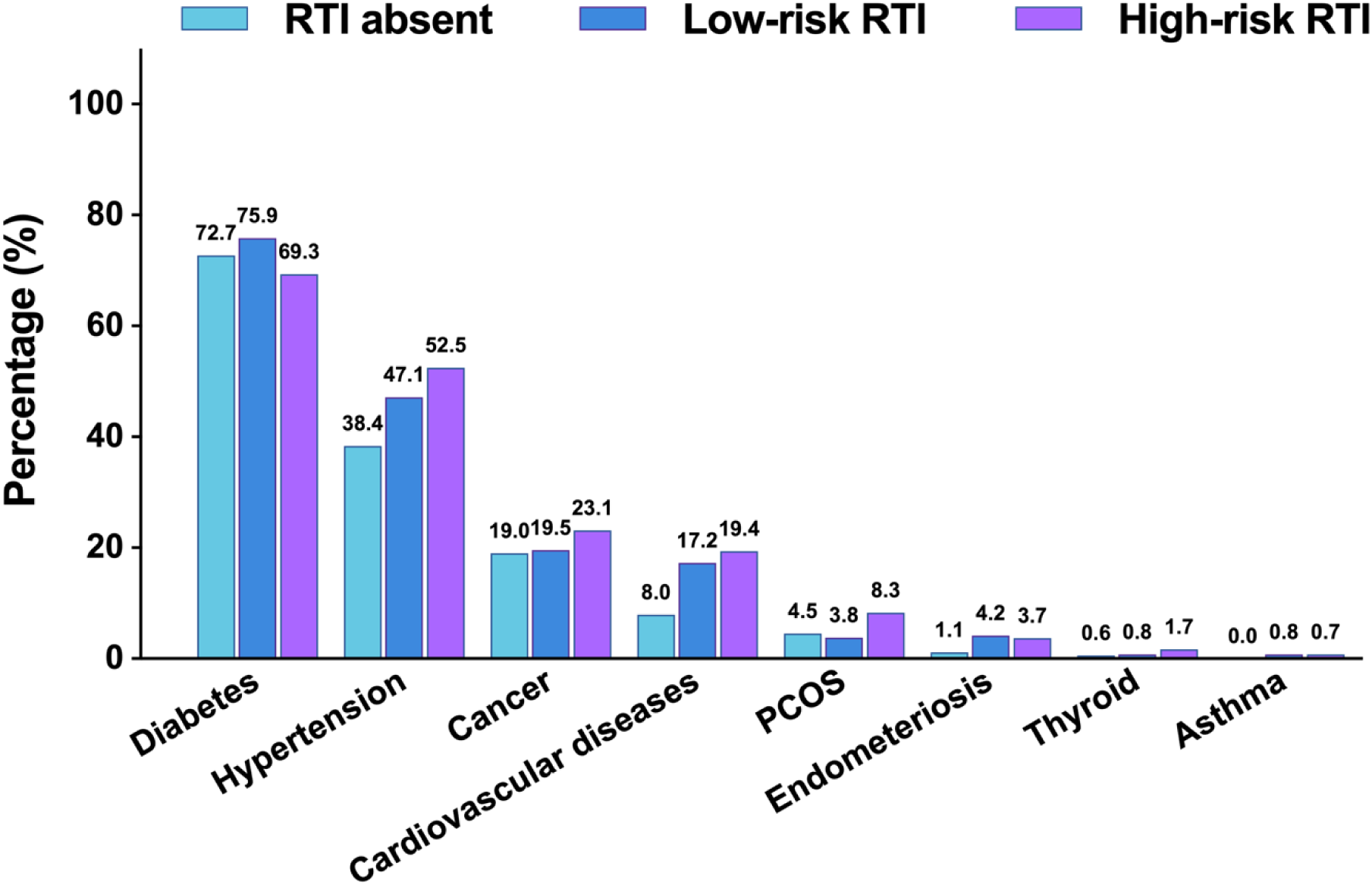
Distribution of Family History of Comorbidities Among Participants. This bar chart illustrates the prevalence of various comorbidities of family members of participants categorized as RTI-absent, low-risk RTI, and high-risk RTI groups. The x-axis represents different comorbid conditions, while the y-axis denotes the percentage of participants with a family history of each condition. RTI, Reproductive Tract Infection; PCOS, Polycystic Ovarian Syndrome. The percentage for each disease is displayed above its corresponding bar in the graph.

**Figure 4.**
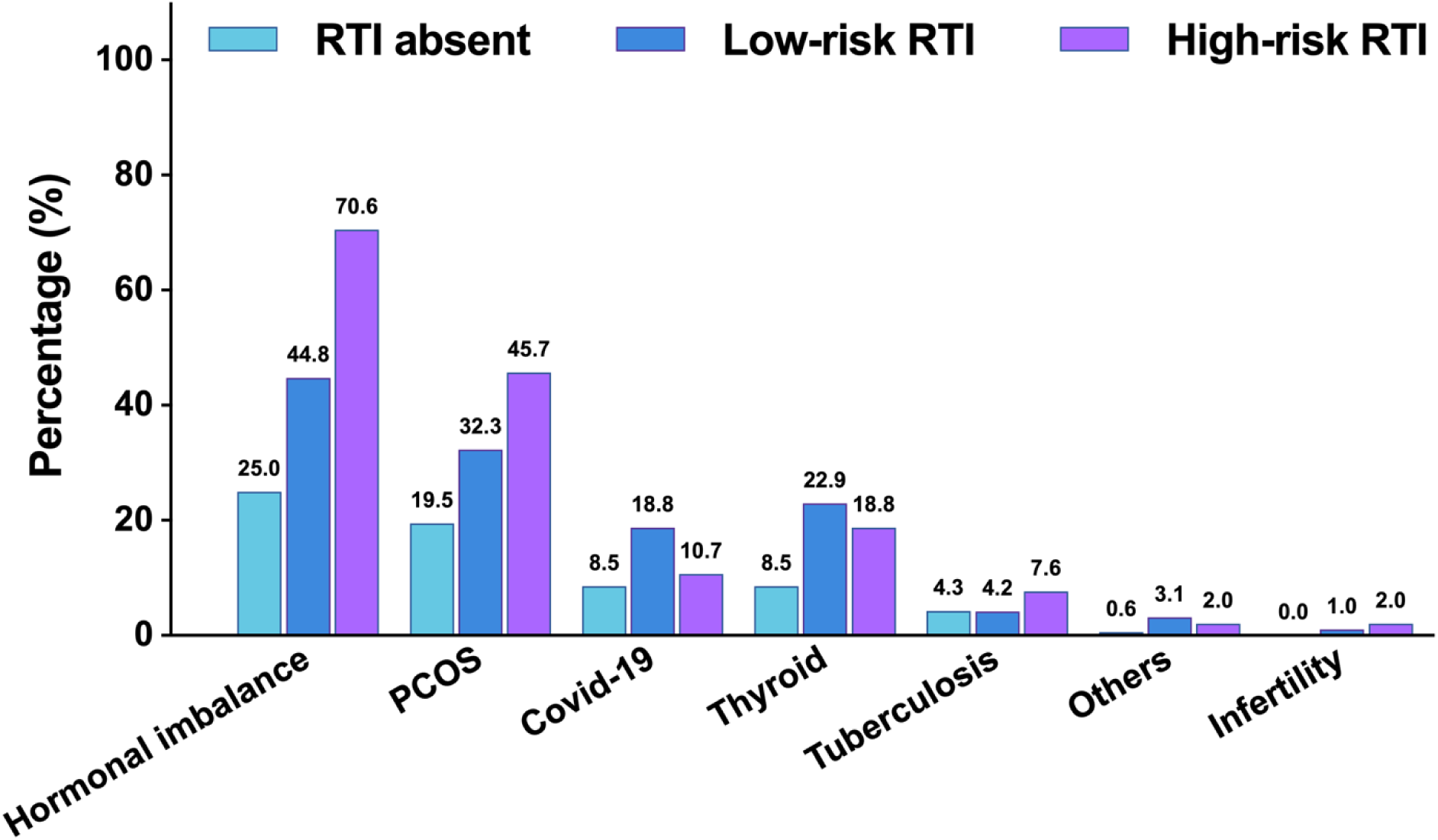
Distribution of Medical Conditions Among RTI-Absent, Low-Risk RTI, and High-Risk RTI Participants. This bar chart presents the prevalence of various medical conditions among participants categorized into RTI-absent, low-risk RTI, and high-risk RTI groups. The x-axis represents different medical conditions, while the y-axis denotes the percentage of participants affected. RTI, Reproductive Tract Infections; PCOS, Polycystic Ovarian Syndrome. The percentage for each disease is displayed above its corresponding bar in the graph.

#### 3.3.4. Vaccines Against RTI

The HPV vaccine uptake was higher in the low-risk RTI group compared to the high-risk and RTI-absent groups (**Table 5**). Additionally, hepatitis B vaccination rates were similar across all three groups.

**Table 5:** Association of HPV and Hepatitis B Vaccination Status with Symptoms Suggestive of Reproductive Tract Infections Among Women.

| Variables | RTI absent<br>N= 679<br>(35.4%) | Low-risk RTI<br>N= 446<br>(23.2%) | High-risk<br>RTI<br>N= 795<br>(41.4%) | Low-risk<br>RTI<br>vs<br>RTI absent<br>$\chi^2$<br>P-value | High-risk<br>RTI<br>vs<br>RTI absent<br>$\chi^2$<br>P-value | Low-risk RTI<br>vs<br>High-risk RTI<br>$\chi^2$<br>P-value |
| --- | --- | --- | --- | --- | --- | --- |
| **HPV Vaccination Status** |  |  |  |  |  |  |
| Yes | 49 (7.2%) | 46 (10.3%) | 63 (7.9%) | 2.9517 | 0.1704 | 1.7486 |
| No | 629 (92.8%) | 400 (89.7%) | 732 (92.1%) | 0.0858 | 0.6798 | 0.1860 |
| **Hepatitis B Vaccination Status** |  |  |  |  |  |  |
| Yes | 161 (23.7%) | 130 (29.1%) | 206 (25.9%) | 3.8145 | 0.8053 | 1.3559 |
| No | 517 (76.3%) | 316 (70.9%) | 589 (74.1%) | 0.0508 | 0.3695 | 0.2443 |

RTI, reproductive tract infections.

### 3.4.. Multinomial Logistic Regression Analysis

Multinomial logistic regression analysis was conducted to identify the risk factors associated with **low- and high-risk RTI** (**Tables 6** and **7**).

#### 3.4.1. Significant Predictors of Participants in the Low-Risk RTI Group

The analysis of the low-risk RTI group identified several significant predictors (**Table 6**). Menstrual health factors were key indicators, with irregular menstrual cycles (*P*=0.035) and prolonged cycle length (*P*=0.040) showing significant associations. Family medical history also exhibited bilateral significance (*P*=0.027, *P*=0.028), suggesting a potential hereditary component in RTI/STI susceptibility. Additionally, contraception awareness (*P*=0.019) was found to be a significant predictor.

**Table 6.** A Multinomial Logistic Regression Analysis of Factors Associated with Low-Risk of Reproductive Tract Infections.

| Variables | OR | 95% CI | P-value |
| --- | --- | --- | --- |
| Age (Years) | 0.1053 | [-3.121, 3.332] | 0.949 |
| BMI status | 0.7879 | [-2.305, 3.881] | 0.618 |
| Life stage | -2.0828 | [-5.662, 1.496] | 0.254 |
| Highest level of education | -0.0633 | [-1.598, 1.471] | 0.936 |
| Occupation | -0.5610 | [-1.426, 0.499] | 0.347 |
| Prescription Medication Usage in the last 3 Months for STI/RTI Symptoms | 0.4602 | [-4.729, 12.191] | 0.080 |
| Age at Menarche (First Period) | 0.2352 | [-2.298, 3.168] | 0.890 |
| Menstrual Cycle Description | -2.3226 | [-4.394, -0.127] | 0.035* |
| Length of Menstrual Cycle | -1.6808 | [-3.289, -0.073] | 0.040* |
| Family History of Medical Conditions | -2.9366 | [-6.593, -0.280] | 0.027* |
| Presence of Chronic Illness | -0.1956 | [-1.548, 1.157] | 0.776 |
| History of Symptoms Related to RTI/STI | 0.3736 | [-2.027, 2.774] | 0.760 |
| Diagnosis of STI/RTI in the Last 3 Months | -57.0043 | [-1.21e+04, 1.51e+04] | 0.998 |
| Menstrual Hygiene Behavior | -2.8428 | [-20.452, 14.767] | 0.752 |
| Awareness of Contraceptive Methods | -2.8564 | [-8.045, -0.888] | 0.019* |
| HPV Vaccination Status | 0.075 | [-0.883, 1.13] | 0.789 |
| Hepatitis B Vaccination Status | 0.023 | [0.205, 0.447] | 0.3065 |

BMI, body mass index; OR, odd ratio; CI, confidence interval; RTI, reproductive tract infection; STI, sexually transmitted infection; \* $P<0.05$ .

#### 3.4.2. Significant Predictors of Participants in the High-Risk RTI Group

The analysis of the high-risk RTI group revealed a distinct pattern of significant predictors (**Table 7**). Medical history played a critical role, with prior RTI/STI medication use showing strong significance (*P*=0.001, OR=5.9358), indicating that a history of treatment substantially increased the likelihood of high-risk classification. Family disease history (*P*=0.033, OR=-4.4745) was also identified as a significant predictor.

**Table 7.** A Multinomial Logistic Regression Analysis of Factors Associated with High-Risk of Reproductive Tract Infections (RTIs)

| Variables | OR | 95% CI | P-value |
| --- | --- | --- | --- |
| Age (Years) | -0.1314 | [-3.486, 3.223] | 0.939 |
| BMI status | 0.3056 | [-2.856, 3.275] | 0.848 |
| Life stage | -2.2586 | [-5.145, 0.622] | 0.125 |
| Highest level of education | -0.3586 | [-1.798, 1.272] | 0.067 |
| Occupation | -0.8635 | [-1.896, 0.269] | 0.131 |
| Prescription Medication Usage in the Last 3 Months for RTI/STI | 5.9358 | [4.183, 13.729] | 0.000* |
| Age at Menarche (First Period) | -2.0336 | [-4.484, 0.117] | 0.064 |
| Menstrual Cycle Description | -1.1638 | [-4.175, 1.836] | 0.446 |
| Length of Menstrual Cycle | -1.0525 | [-3.258, 0.064] | 0.064 |
| Family History of Medical Conditions | -4.4745 | [-6.728, -0.198] | 0.033* |
| Presence of Chronic Illness | -0.8130 | [-2.680, 2.054] | 0.900 |
| History of Symptoms Related to RTI/STI | -0.6116 | [-3.031, 2.168] | 0.745 |
| Diagnosis of STI/RTI in the Last 3 Months | 16.3334 | [-1.51e+04, 1.51e+04] | 0.999 |
| Menstrual Hygiene Behavior | -2.3624 | [-4.462, -0.263] | 0.027* |
| Awareness of Contraceptive Methods | -0.9448 | [-8.116, -0.789] | 0.003* |
| HPV Vaccination Status | 0.089 | [-0.983, 1.23] | 0.889 |
| Hepatitis B Vaccination Status | -0.523 | [-1.205, 0.447] | 0.206 |

BMI, body mass index; OR, Odd Ratio; CI, confidence interval; RTI, reproductive tract infection; STI sexually transmitted infection; \* $P < 0.05$ .

Additionally, menstrual hygiene behavior (*P*=0.027, OR=-2.3624) and contraception awareness (*P*=0.003, OR=-0.9448) emerged as key factors associated with high-risk RTI classification.

### 3.5. Acceptance and Hesitancy Toward RTI/STI Vaccination

Participants were asked, “If available today, would you be interested in receiving a vaccine to prevent STIs/RTIs?” Among the 1,920 participants, 46.2% (N=888) expressed willingness to receive an STI/RTI vaccine, indicating vaccine acceptance, while 53.8% (N=1,032) were not interested, reflecting vaccine hesitancy.

When asked, “If and when available, which of the following STI vaccines would you be interested in receiving?”, 56.8% (*N*=508) of vaccine-accepting participants responded. Among them, 96.1% showed interest in an HIV vaccine, followed by herpes (60.4%), syphilis (56.8%), gonorrhea (55.8%), chlamydia (47.1%), and trichomoniasis (43.6%) (**Figure 5**).

**Figure 5.**
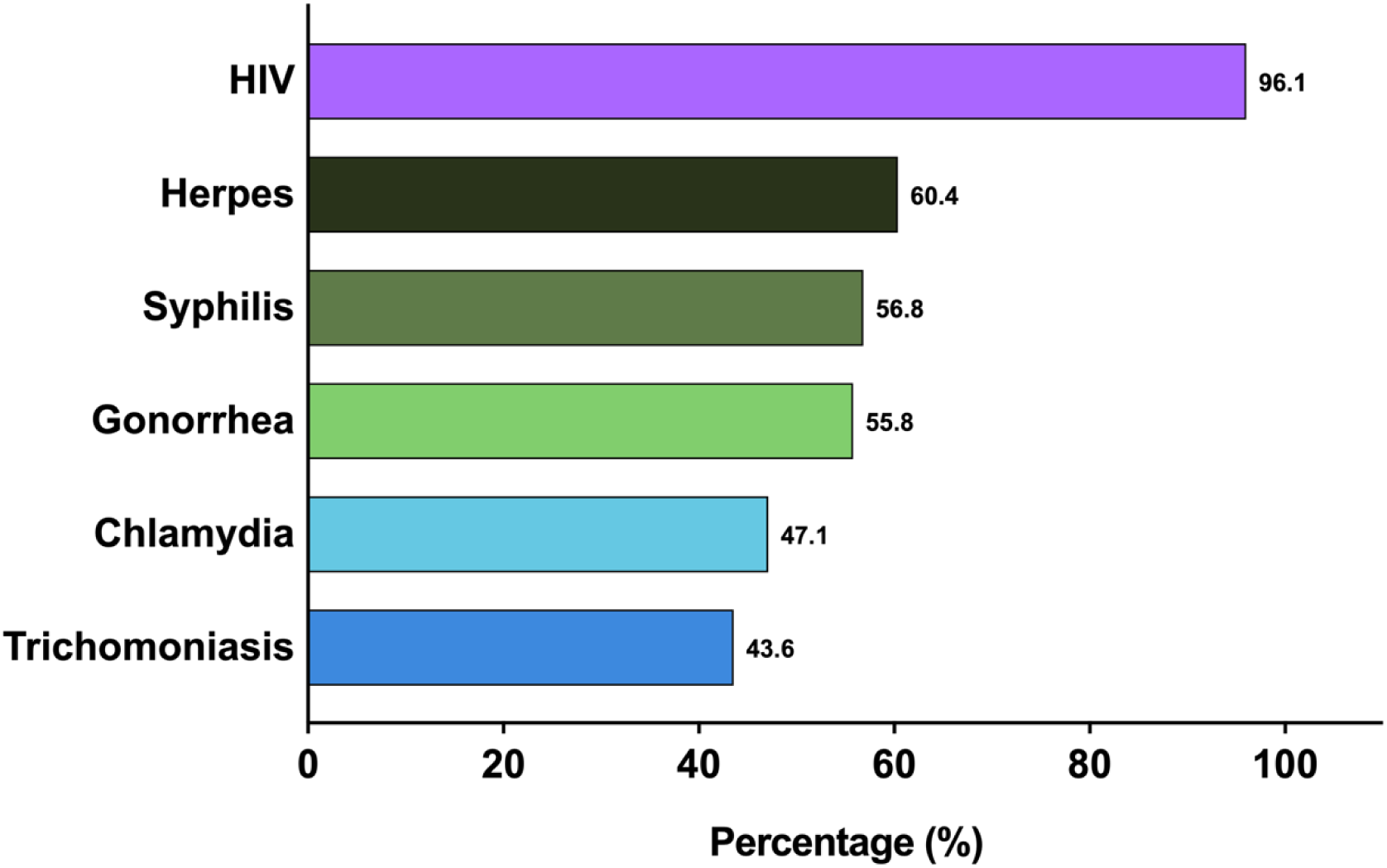
Distribution of Diseases for Which Participants Expressed Interest in Vaccination. This horizontal bar chart presents the percentage of participants interested in receiving vaccines for sexually transmitted infections (STIs). The x-axis represents the percentage of participants, while the y-axis lists the specific diseases for which vaccination interest was recorded.

Additionally, participants were asked, “If STI vaccines were available, when do you think it would be best to first offer STI vaccines?” The majority (50.5%) believed adolescence was the ideal time, followed by early adulthood (23.5%) (**Table 1**).

### 3.6 Factors Influencing RTI/STI Vaccine Acceptance and Hesitancy Among Urban Women

#### 3.6.1. Socio-Demographic Factors

Participants were categorized into two groups based on their intention to receive an RTI/STI vaccine: the vaccine acceptance group and the vaccine-hesitant group (**Table 8**). The Chi-square test (*χ²*) was used to assess demographic differences between these groups.

**Table 8.**
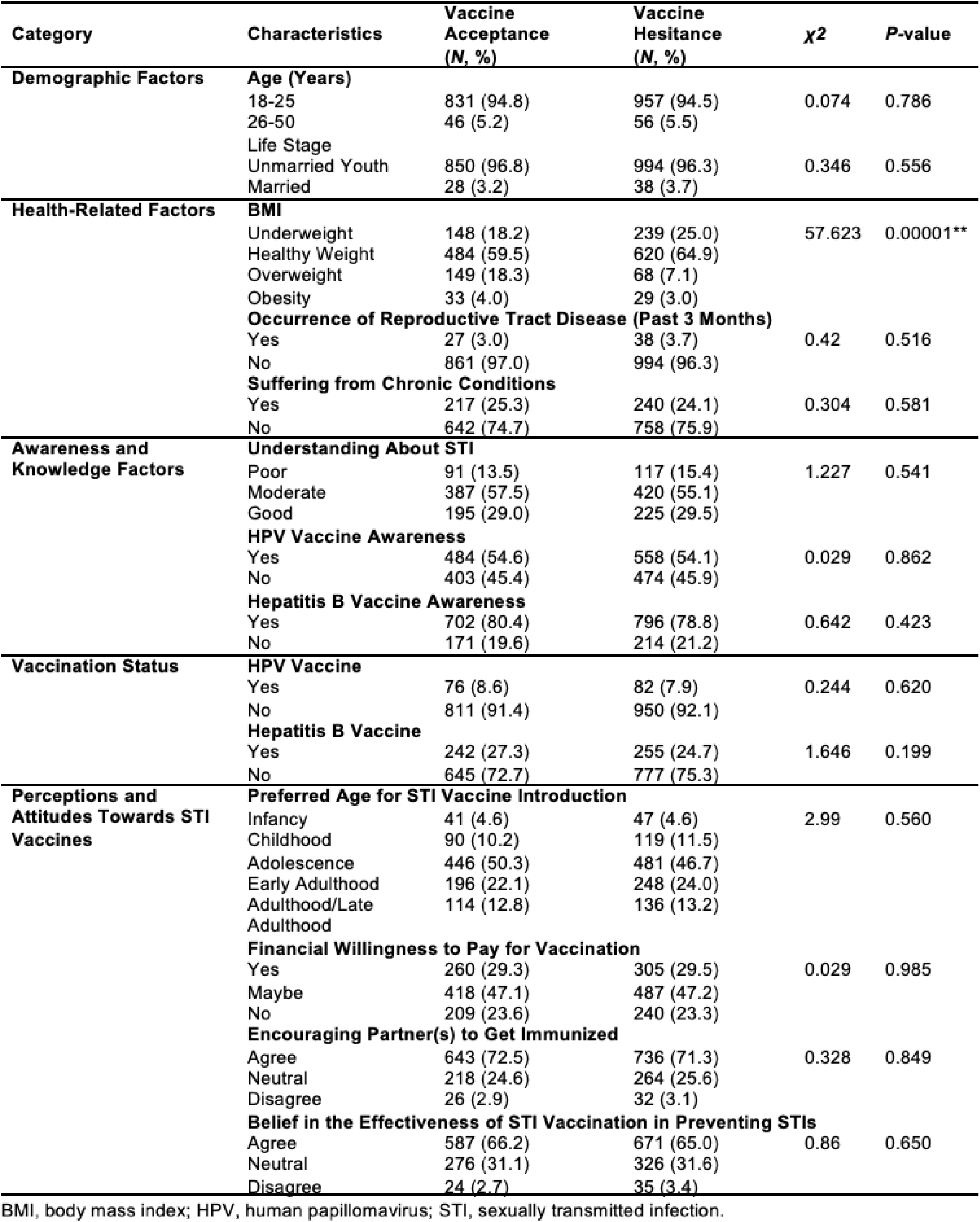
Comparison of Demographic, Health, and Awareness Factors Between Vaccine Acceptance (7^888, 46.2%) and Hesitancy Groups (^1032, 53.8%)

| Category | Characteristics | Vaccine Acceptance (N, %) | Vaccine Hesitancy (N, %) | $\chi^2$ | P-value |
| --- | --- | --- | --- | --- | --- |
| **Demographic Factors** | **Age (Years)** |  |  |  |  |
|  | 18-25 | 831 (94.8) | 957 (94.5) | 0.074 | 0.786 |
|  | 26-50 | 46 (5.2) | 56 (5.5) |  |  |
|  | **Life Stage** |  |  |  |  |
|  | Unmarried Youth | 850 (96.8) | 994 (96.3) | 0.346 | 0.556 |
|  | Married | 28 (3.2) | 38 (3.7) |  |  |
| **Health-Related Factors** | **BMI** |  |  |  |  |
|  | Underweight | 148 (18.2) | 239 (25.0) | 57.623 | 0.00001** |
|  | Healthy Weight | 484 (59.5) | 620 (64.9) |  |  |
|  | Overweight | 149 (18.3) | 68 (7.1) |  |  |
|  | Obesity | 33 (4.0) | 29 (3.0) |  |  |
|  | **Occurrence of Reproductive Tract Disease (Past 3 Months)** |  |  |  |  |
|  | Yes | 27 (3.0) | 38 (3.7) | 0.42 | 0.516 |
|  | No | 861 (97.0) | 994 (96.3) |  |  |
|  | **Suffering from Chronic Conditions** |  |  |  |  |
|  | Yes | 217 (25.3) | 240 (24.1) | 0.304 | 0.581 |
|  | No | 642 (74.7) | 758 (75.9) |  |  |
| **Awareness and Knowledge Factors** | **Understanding About STI** |  |  |  |  |
|  | Poor | 91 (13.5) | 117 (15.4) | 1.227 | 0.541 |
|  | Moderate | 387 (57.5) | 420 (55.1) |  |  |
|  | Good | 195 (29.0) | 225 (29.5) |  |  |
|  | **HPV Vaccine Awareness** |  |  |  |  |
|  | Yes | 484 (54.6) | 558 (54.1) | 0.029 | 0.862 |
|  | No | 403 (45.4) | 474 (45.9) |  |  |
|  | **Hepatitis B Vaccine Awareness** |  |  |  |  |
|  | Yes | 702 (80.4) | 796 (78.8) | 0.642 | 0.423 |
|  | No | 171 (19.6) | 214 (21.2) |  |  |
| **Vaccination Status** | **HPV Vaccine** |  |  |  |  |
|  | Yes | 76 (8.6) | 82 (7.9) | 0.244 | 0.620 |
|  | No | 811 (91.4) | 950 (92.1) |  |  |
|  | **Hepatitis B Vaccine** |  |  |  |  |
|  | Yes | 242 (27.3) | 255 (24.7) | 1.646 | 0.199 |
|  | No | 645 (72.7) | 777 (75.3) |  |  |
| **Perceptions and Attitudes Towards STI Vaccines** | **Preferred Age for STI Vaccine Introduction** |  |  |  |  |
|  | Infancy | 41 (4.6) | 47 (4.6) | 2.99 | 0.560 |
|  | Childhood | 90 (10.2) | 119 (11.5) |  |  |
|  | Adolescence | 446 (50.3) | 481 (46.7) |  |  |
|  | Early Adulthood | 196 (22.1) | 248 (24.0) |  |  |
|  | Adulthood/Late Adulthood | 114 (12.8) | 136 (13.2) |  |  |
|  | **Financial Willingness to Pay for Vaccination** |  |  |  |  |
|  | Yes | 260 (29.3) | 305 (29.5) | 0.029 | 0.985 |
|  | Maybe | 418 (47.1) | 487 (47.2) |  |  |
|  | No | 209 (23.6) | 240 (23.3) |  |  |
|  | **Encouraging Partner(s) to Get Immunized** |  |  |  |  |
|  | Agree | 643 (72.5) | 736 (71.3) | 0.328 | 0.849 |
|  | Neutral | 218 (24.6) | 264 (25.6) |  |  |
|  | Disagree | 26 (2.9) | 32 (3.1) |  |  |
|  | **Belief in the Effectiveness of STI Vaccination in Preventing STIs** |  |  |  |  |
|  | Agree | 587 (66.2) | 671 (65.0) | 0.86 | 0.650 |
|  | Neutral | 276 (31.1) | 326 (31.6) |  |  |
|  | Disagree | 24 (2.7) | 35 (3.4) |  |  |

BMI, body mass index; HPV, human papillomavirus; STI, sexually transmitted infection.

Most characteristics, including age, life stage, occupation, education status, RTI occurrence, and chronic conditions, were similar between the two groups. However, BMI (*χ²* = 57.62, *P*<0.0001) showed a statistically significant difference.

The percentage of underweight participants was higher in the vaccine-hesitant group (25.0%) compared to the vaccine acceptance group (18.2%).

#### 3.6.2. Knowledge and Awareness of STIs and STI Vaccination Among Vaccine-Accepting and Vaccine-Hesitant Participants

Participants were asked about their understanding of STIs, and no significant differences were observed between the vaccine acceptance and vaccine-hesitant groups. However, the majority in both groups demonstrated moderate knowledge (57.5% vs. 55.1%) (**Table 8**).

Additionally, participants were assessed on their awareness of STI vaccines, specifically HPV and hepatitis B. The results indicated that both groups had nearly equal levels of awareness about these vaccines (**Table 8**). However, a significantly higher percentage of participants (∼80%) were aware of the hepatitis B vaccine, compared to ∼54% for the HPV vaccine.

Similarly, the percentage of participants who had received the hepatitis B vaccine (27.3%) was notably higher than those who had received the HPV vaccine (8.6%). However, no significant differences were observed in the distribution of HPV and hepatitis B vaccination rates between the vaccine acceptance and vaccine-hesitant groups.

#### 3.6.3. Financial Considerations and Perceived Effectiveness of STI Vaccination

Approximately ∼29% of participants from both the vaccine acceptance and vaccine-hesitant groups were willing to pay for an STI vaccine. However, 47% remained uncertain, selecting the response “maybe.”

When asked, “If an STI vaccine were available, would you encourage your partner(s) to get immunized?”, more than half of the participants in both groups expressed support for their partners getting vaccinated (**Table 8**).

Participants were also asked, “Do you think STI vaccination could be an effective way to prevent STIs?” No significant differences were observed between the vaccine acceptance and vaccine-hesitant groups regarding their belief in vaccine effectiveness. Notably, even among those hesitant to receive the vaccine, 65% still believed in its effectiveness. Given this finding, we further examined barriers to STI vaccination (**Figure 6**).

**Figure 6.**
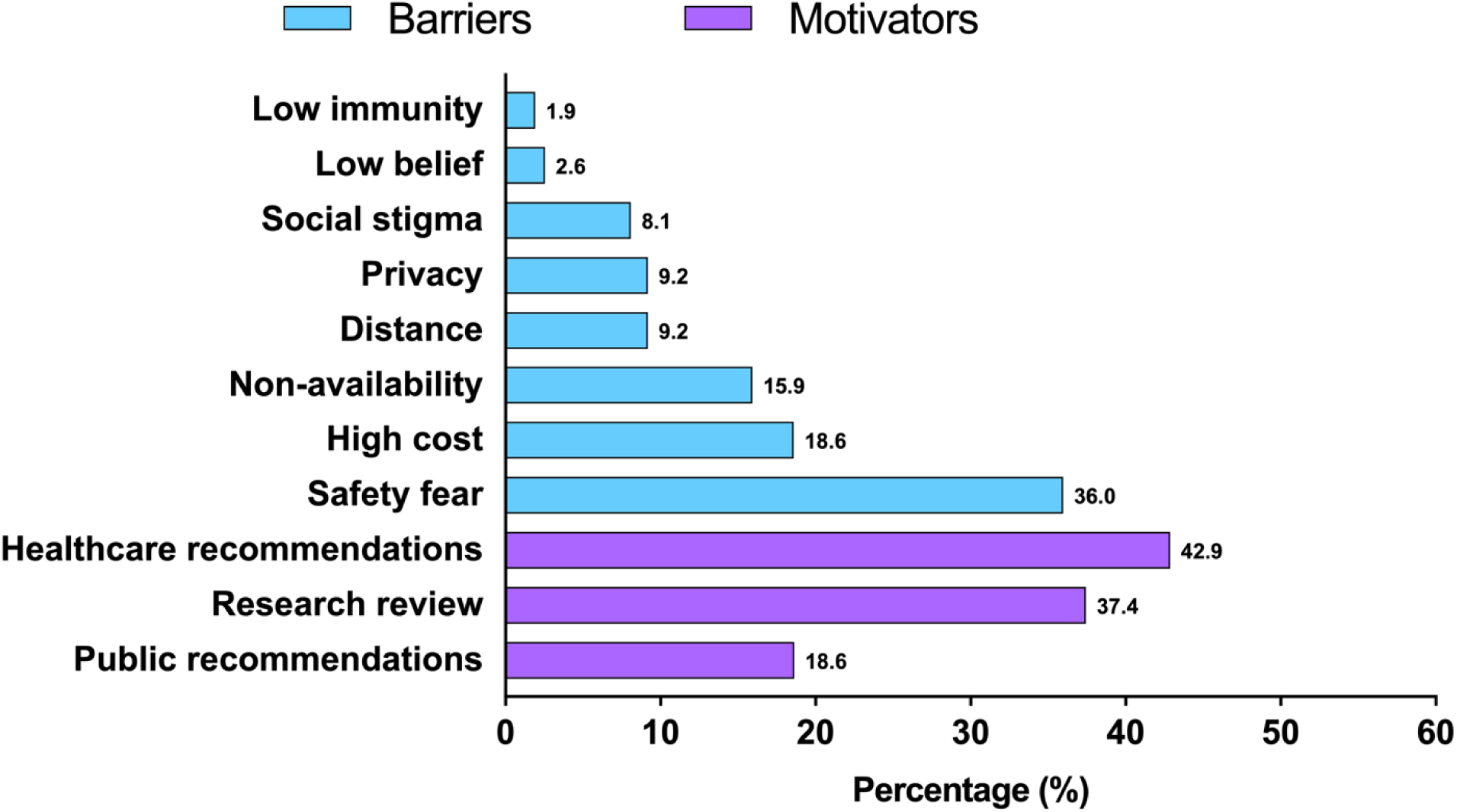
Barriers and Motivators Shaping Interest in Sexually Transmitted Infection (STI) Vaccination. This figure presents the distribution of barriers (blue) and motivating factors (purple) associated with individuals’ reluctance or willingness or to receive an STI vaccine. The x-axis represents the percentage of respondents reporting each factor. The percentage for each source is displayed above its corresponding bar in the graph.

#### 3.6.4. Barriers to STI Vaccine Acceptance

Participants reported various barriers to receiving an STI vaccine (**Figure 6**). The most common concern was vaccine safety (36%), followed by cost concerns (18.6%).

Other reported barriers included vaccine non-availability (15.9%) and the inconvenience of accessing a clinic due to distance (9.2%). Additionally, privacy concerns were a factor, with 9.2% of participants worried about having STI vaccination recorded in their health records and 8.1% citing social stigma as a deterrent.

The least prevalent barriers were trust-related concerns, such as low belief in vaccines (2.6%) and health-related concerns including low immunity, older age, or comorbidities (1.9%).

#### 3.6.5. Prevalence of Motivating Factors for STI Vaccine Acceptance Among the Hesitant Group

Among participants who answered the question, “Which of the following would be helpful in accepting an STI vaccine?”, the most common motivating factor was a recommendation from a doctor or healthcare provider (∼43%) (**Figure 6**).

Additionally, 37.4% of participants stated they would be motivated after reviewing detailed research on vaccine adverse events and its constituents. A smaller yet notable proportion (18.6%) expressed concerns about vaccine safety, indicating they would wait to observe others’ reactions before making a decision.

#### 3.6.6. Preferred and Trusted Sources of Information on STI Vaccines

Participants were asked, “Where would you like to receive information about STI vaccines?” As shown in **Figure 7**, the most trusted source was healthcare providers (48.7%), followed by reliable organizations (32.8%), such as University of British Columbia (UBC), National Institutes of Health (NIH), Centers for Disease Control and Prevention (CDC), and WHO.

**Figure 7.**
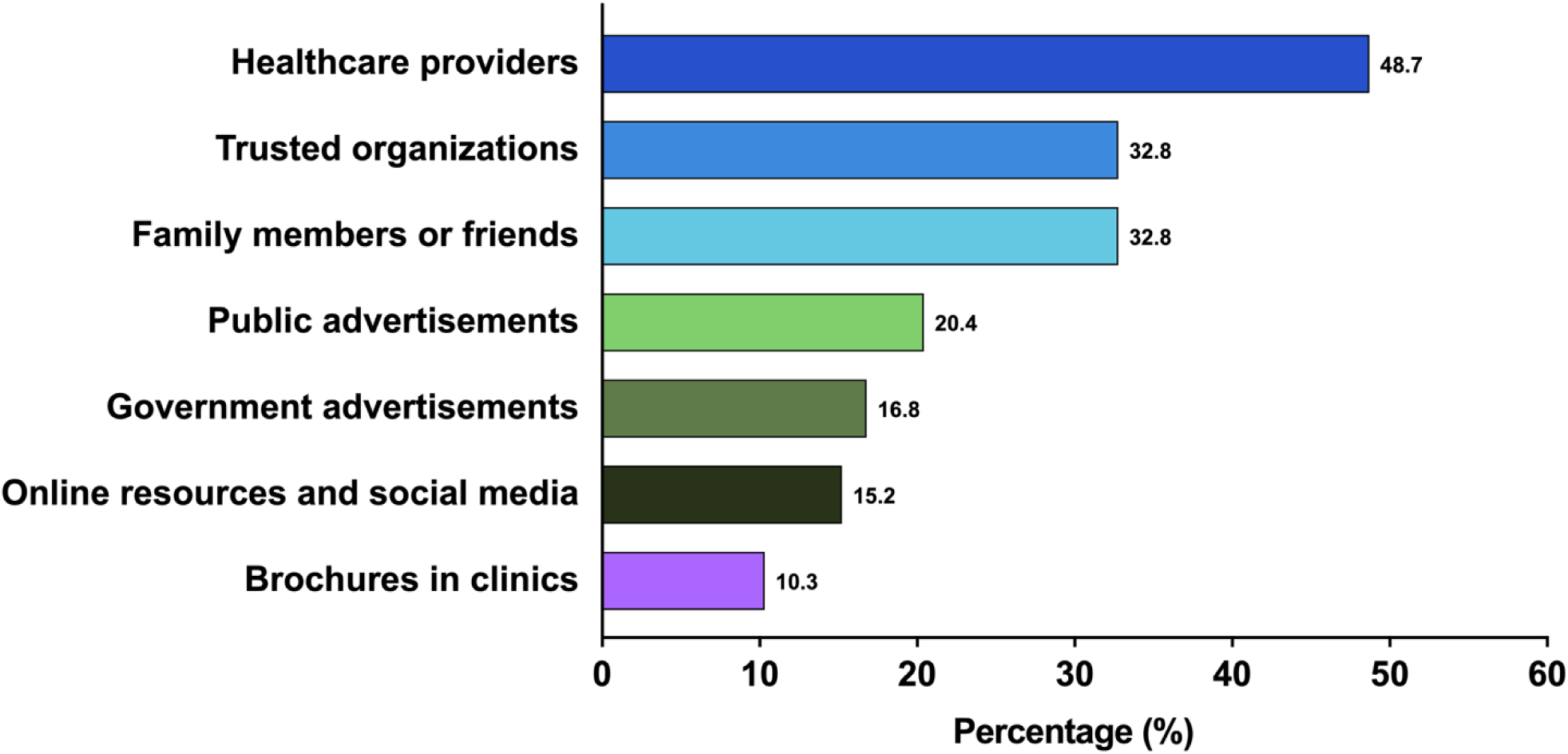
Trusted Sources of Information About Sexually Transmitted Infection (STI) Vaccination. This figure illustrates the distribution of trusted sources from which individuals seek information regarding STI vaccines. The x-axis represents the percentage of respondents who consider each source reliable.

Other sources included family members and friends (32.8%), public advertisements (20.4%), and government advertisements (16.8%). Additionally, 15.2% of participants reported relying on online resources and social media, while 10.3% obtained information from brochures in clinics.

## 4. Discussion

This study aimed to determine the prevalence and risk factors of reproductive tract infections (RTIs) among urban women in Delhi/NCR. The findings reveal a substantial burden of RTI symptoms, with ∼65% of participants overall and ∼41% classified as high-risk. These symptoms are commonly linked to RTI such as bacterial vaginosis and candidiasis. Abnormal vaginal discharge was the most frequently reported symptom, aligning with previous studies [19, 20]. Additionally, abdominal pain and backache, experienced by more than 65% of participants, were consistent with prior findings among North Indian women [21, 22]. Despite the high prevalence of symptoms, only 5% of participants reported a gynecological diagnosis of RTI/STI, suggesting significant underdiagnosis and underreporting. Factors contributing to this discrepancy may include social stigma, lack of awareness, and limited access to healthcare services. Pelvic inflammatory disease (39.5%) was the most common gynecological condition in both low- and high-risk RTI groups, while vaginitis (37.2%) was more prevalent in the high-risk RTI group. These findings align with previous research [23] and underscore the importance of regular screening and comprehensive treatment plans to effectively manage RTI. Interestingly, unlike previous studies that identified age, marital status, and education as critical socio-demographic determinants of RTI risk [23, 24], the present study found no significant association with these variables. This lack of variability could be attributed to the predominant participation of urban women, which may have influenced the findings.

For the first time, this study identified a significant association between family comorbidities, particularly diabetes and hypertension, and high-risk RTI. A strong link with familial medical history suggests that genetic predisposition to diabetes may impair immune responses, increasing susceptibility to infections. These observations emphasize the importance of addressing comorbid conditions in reproductive health management. Menstrual health factors also played a crucial role in RTI risk. Women with menstrual cycles longer than 45 days had a significantly higher likelihood of high-risk RTI. Additionally, hormonal imbalances, particularly PCOS, were more prevalent in the high-risk RTI group, reinforcing the role of hormonally mediated immune responses in increasing susceptibility to infections. These results align with previous research [25, 26], which links irregular cycles and PCOS to RTI [27, 28]. Regular monitoring of menstrual irregularities and timely PCOS management could help mitigate RTI risk. Another potential risk factor was early menarche. Women who experienced menarche between ages 10–12 were more likely to report both low- and high-risk RTI compared to those with menarche onset at 12–16 years or older. Early menarche is a known risk factor for reproductive health issues, likely due to prolonged estrogen exposure and early sexual initiation [29, 30]. Poor menstrual hygiene emerged as another significant risk factor for high-risk RTI, reinforcing previous findings that inadequate hygiene facilitates pathogen growth and infections [31, 32]. Contrary to some studies, this research found that women with low- and high-risk RTI exhibited higher awareness of contraceptive methods [33]. However, this may indicate that these groups engage in higher sexual activity, increasing infection risk [19].

A history of RTIs/STIs was significantly associated with high-risk RTI, suggesting that recurrent or untreated infections contribute to reproductive health deterioration. A considerable proportion of participants (35.0% in the low-risk and 30.3% in the high-risk groups) had taken medication for RTIs/UTIs symptoms within the past three months, indicating a persistent burden of symptomatic RTI even among low-risk groups. Previous studies have also reported recurrent RTI in prior patients [22, 31]. However, the continued presence of symptoms raises concerns about medication adherence, incomplete treatment, or misdiagnosis.

In addition to RTI prevalence and risk factors, this study examined urban women’s willingness to receive STIs/RTIs vaccinations. Consistent with prior research, low uptake of HPV and hepatitis B vaccines remains a significant public health challenge, particularly in low- and middle-income countries [34]. While most HPV vaccinations occurred during adolescence (ages 11–18), in line with global immunization guidelines, hepatitis B vaccinations were primarily received during infancy, reflecting successful early immunization programs [35–37]. However, vaccination efforts for older populations remain inadequate, highlighting the need for targeted awareness campaigns. This study found a moderate vaccine acceptance rate (46.2%), with a high preference for vaccines against HIV, herpes, and syphilis, indicating awareness of infection severity and a willingness to seek protection. The preference for adolescent vaccination (50.5%) aligns with WHO recommendations for early immunization [36, 38]. While overall STI knowledge was similar between the vaccine-accepting and vaccine-hesitant groups, a higher percentage of participants were aware of the hepatitis B vaccine compared to HPV, suggesting that targeted educational efforts could improve HPV vaccine uptake, which remains below average despite its proven efficacy [39]. Vaccine cost was a significant determinant of STI vaccine acceptance, indicating that financial barriers contribute to vaccine hesitancy, particularly in developing countries [40]. However, a positive finding was that more than 50% of participants, regardless of vaccine acceptance status, were willing to encourage their partners to get immunized, highlighting a potential community health benefit. Although vaccine hesitancy was prevalent, 65% of participants believed in vaccine efficacy, suggesting that factors beyond efficacy—such as cost and accessibility—contribute to hesitancy. Vaccine safety concerns were the most cited barrier, followed by cost, availability, and access [41]. Social stigma surrounding STI vaccines also emerged as a major issue, emphasizing the need for sensitization efforts to improve acceptance [42].

Interestingly, attitudinal barriers such as low belief in vaccines and immunity concerns were less common, suggesting that misconceptions about vaccine effectiveness are not the main drivers of hesitancy. The strongest motivator for vaccine acceptance was a recommendation from a healthcare provider, followed by a detailed research review, reinforcing previous findings that healthcare professionals play a crucial role in addressing vaccine concerns [43, 44]. Similarly, healthcare providers were the most trusted source of information, followed by WHO, CDC, family and friends, and public advertisements [16, 45].

## 5. Conclusion

This study highlights a high prevalence of symptomatic RTIs among urban women in Delhi/NCR, with key contributing factors including family history of comorbidities, early menarche, irregular menstrual cycles, and inadequate menstrual hygiene. Vaccine hesitancy, primarily driven by safety and cost concerns, underscores the need for enhanced awareness, improved access, and stronger preventive healthcare measures.

Given the established link between RTI and increased HIV susceptibility, these findings further reinforce the urgent need for integrated STI and HIV prevention strategies. Improving screening programs, expanding STI vaccine coverage, and addressing social stigma could significantly reduce the dual burden of RTI and HIV, particularly in resource-limited settings. Additionally, future vaccine development efforts, including HIV and multi-pathogen STI vaccines, should consider these findings to enhance public acceptance and uptake. Strengthening healthcare infrastructure, provider recommendations, and targeted awareness campaigns remains essential to mitigate RTI/STI-associated risks and reduce overall infection rates.

## 6. Limitations of the Study

This study did not include laboratory tests for diagnosing RTI/STIs and instead relied on a syndromic approach, which may not accurately estimate the true prevalence of infections in the community. Additionally, the syndromic approach may have excluded asymptomatic RTI cases, potentially leading to an underestimation of the actual disease burden.

Social stigma surrounding RTI may have also discouraged some women from reporting their symptoms, further contributing to underreported prevalence rates. Lastly, this study did not include follow-up with participants who sought or were encouraged to seek treatment, limiting insights into treatment adherence and long-term health outcomes.

## Author Contribution

Conceptualization, J.T. and A.E.S.; Questionnaire designing, J.T., A.E.S., S.K.Y., R.K., D.S., S.M. and R.S.; Data collection, J.T., S.K.Y., P.B. and A.G.M.; Data Analysis, J.T., P.B., S.K.Y., R.K. and J.G.; Writing – Original Draft Preparation, S.K.Y. and P.B.; Review & Editing, J.T., A.E.S., R.K. and S.M.; Supervision, J.T. and A.E.S.; Visualization, J.T. and A.E.S.; Project Administration, J.T. and A.E.S.; Funding Acquisition, A.E.S.

## Institutional Review Board Statement

The study has been approved by Institutional Human Ethics Committee at Dr. B.R. Ambedkar Center for Biomedical Research(ACBR), University of Delhi, Delhi, India.

## Informed Consent Statement

Informed consent was obtained from all subjects involved in the study.

## Data Availability Statement

All data will be available on request from the corresponding authors.

## Acknowledgments

We thank Dr. Mamtesh Singh, Dr. Manisha Arora Pandit, Dr. Anshu Arora Anand, Dr. Shivani Kumari, Suman Yadav, and all the students for their assistance in data collection. We also extend our gratitude to Prof. Savita Roy (Principal, Daulat Ram College, University of Delhi) for her logistical support and cooperation.

## Appendix A

**Supplementary Figure S1.**
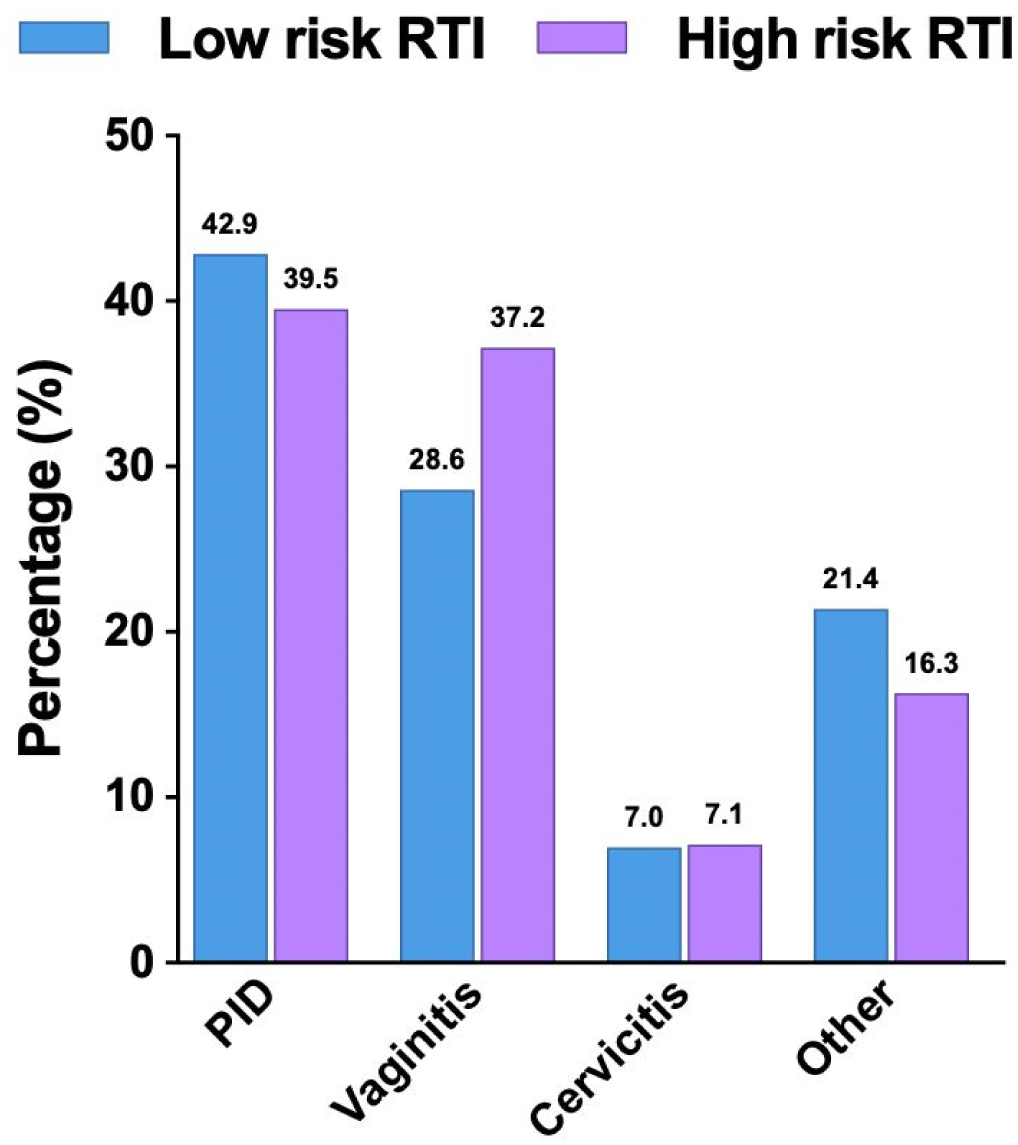
Distribution of Diagnosed RTI Among Low- and High-Risk RTI Participants. This bar chart presents the prevalence of diagnosed gynecological condition associated with reproductive tract infections (RTI) among participants classified as low-risk and high-risk. The x-axis represents different gynecological condition, while the y-axis shows the percentage of participants experiencing each condition. PID; pelvic inflammatory disease

